# Estimation of GP visits, hospitalizations and deaths attributable to RSV and influenza and costs associated with hospitalizations in older adults in France, 2010-2020

**DOI:** 10.1101/2025.04.29.25326541

**Authors:** Charles Nuttens, Vanessa Barbet, Clélia Bignon-Favary, Emilie Lambourg, Stéphane Fiévez, Emmanuelle Blanc, Matias Vacheret, Hervé Liliu, Philippe Vanhems, Jean-Sébastien Casalegno, Laurence Watier, Paul Loubet, Caihua Liang, Elizabeth Begier, Magali Lemaitre

## Abstract

**Background:** Respiratory syncytial virus (RSV) is a leading cause of respiratory infection, causing substantial numbers of hospitalizations and deaths, particularly among vulnerable groups such as infants and older adults. However, its true burden in the adult population is often underestimated due to diagnostic challenges and infrequent standard-of-care testing. This study aimed to estimate the incidence of general practitioner (GP) visits, hospitalizations, and deaths attributable to RSV in French adults aged ≥65 years using a time-series model-based approach. In addition, costs associated with hospitalizations were calculated.

**Methods:** Cyclic Poisson regression models and weekly data from French medical administrative databases and electronic medical records over ten epidemic seasons (2010–2020) were used to estimate incidences for RSV and influenza. The results were stratified by age group, diagnosis causes (respiratory and cardiorespiratory) and diagnosis type (primary and secondary). Average costs per hospitalization were calculated and multiplied by the number of hospitalizations estimated.

**Results:** Among adults aged ≥65 years, we estimated RSV infection was responsible for 647,619 GP visits 24,319 hospitalizations, and 878 deaths per year. Incidence rates for GP visits for RSV were twice as large as for influenza; hospitalization rates were similar and mortality was lower. The mean annual cost of RSV-attributable hospitalizations was 105 million €, similar to influenza.

**Conclusions:** This study highlighted the burden of RSV disease in the adult population in France is higher than previous reported. We envisage that this model-based approach will be instrumental in evaluating the impact of RSV vaccination campaigns.

**Key points:**

- The estimated incidence of RSV infection in older adult is higher than previously reported.
- RSV is responsible for more GP visits, similar hospitalisations but fewer deaths compared to influenza.
- Model-based studies are essential to study RSV epidemiology.

## INTRODUCTION

Respiratory syncytial virus (RSV) infection typically causes mild illness, characterized by flu-like symptoms such as a cough, rhinorrhoea, and wheezing. However, it can also lead to severe respiratory complications, decompensation of underlying diseases, and even death in vulnerable populations such as infants, older adults, immunocompromised individuals, and patients with comorbidities(1,2). While extensively studied in paediatric populations, the burden of RSV infection in adults remains under-recognized (3).

Model-based approaches commonly used for the study of influenza epidemiology are increasingly applied in RSV research (4,5). A recent systematic literature review, meta-analysis, and modelling study estimated a pooled annual incidence of RSV-related hospitalization in adults aged ≥65 years in high-income countries at 157 per 100,000 persons, with a corrected estimate of 347 per 100,000 when accounting for diagnostic limitations (e.g., low sensitivity of single specimen testing, and differences in test type such as DFA vs PCR) (6). These models rely on time-series analysis comparing weekly variations in hospitalizations or deaths and viral activity to estimate the proportion of hospitalizations or deaths attributable to RSV (7).

A recent analysis of the French national hospital database using RSV-specific ICD-10 codes in adults aged ≥18 years reported an incidence rate of 7.2 hospitalizations due to RSV per 100,000 adults aged ≥18 years per year between 2012 and 2021(8). This incidence rate is likely underestimated due to the non-specific symptoms of RSV infection, infrequent standard-of-care testing, low diagnostic accuracy, and the misuse of RSV-specific ICD-10 codes (6,9–15).

The primary study objective was to estimate the incidence of general practitioner (GP) visits, hospitalizations, and deaths attributable to RSV in adults aged ≥65 using multiple time-series regression modelling. We also compared RSV and influenza estimated incidence, examined incidence by age group from 50 years, and characterized hospitalizations and calculated related costs.

## METHODS

### Study design

This is a model-based study using general population data to estimate the annual number of GP visits, hospitalizations, and deaths attributable to RSV and influenza, and the rates per 100,000 population among French adults aged ≥65 years and for the age groups ≥50-64, 65-74, and ≥75. Costs of RSV- and influenza-associated hospitalizations were calculated by the average cost of hospitalization for each incidence rate estimate.

### Data sources

#### General practitioner visits

Weekly numbers of GP visits for respiratory (ICD-10 codes: J00-J99) and cardiorespiratory (ICD-10 codes: I21, I50, I63, I64, J00–J99) causes, stratified by age group, were obtained from the GP network Electronic Medical Records (EMR) database (Figure 1, Supplementary Figure 1). Cardiac codes here and for further indicators were selected based on their associations with cardiac manifestations of RSV and influenza infection (Supplementary Table 1) (16–18). The EMR database collects electronic medical records from 1,200 GPs in France (representing approximately 2% of all French GPs). The panel of contributing GPs is representative of the primary care physician population, based on three main criteria: age, sex, and geographical region.

**Figure 1.**
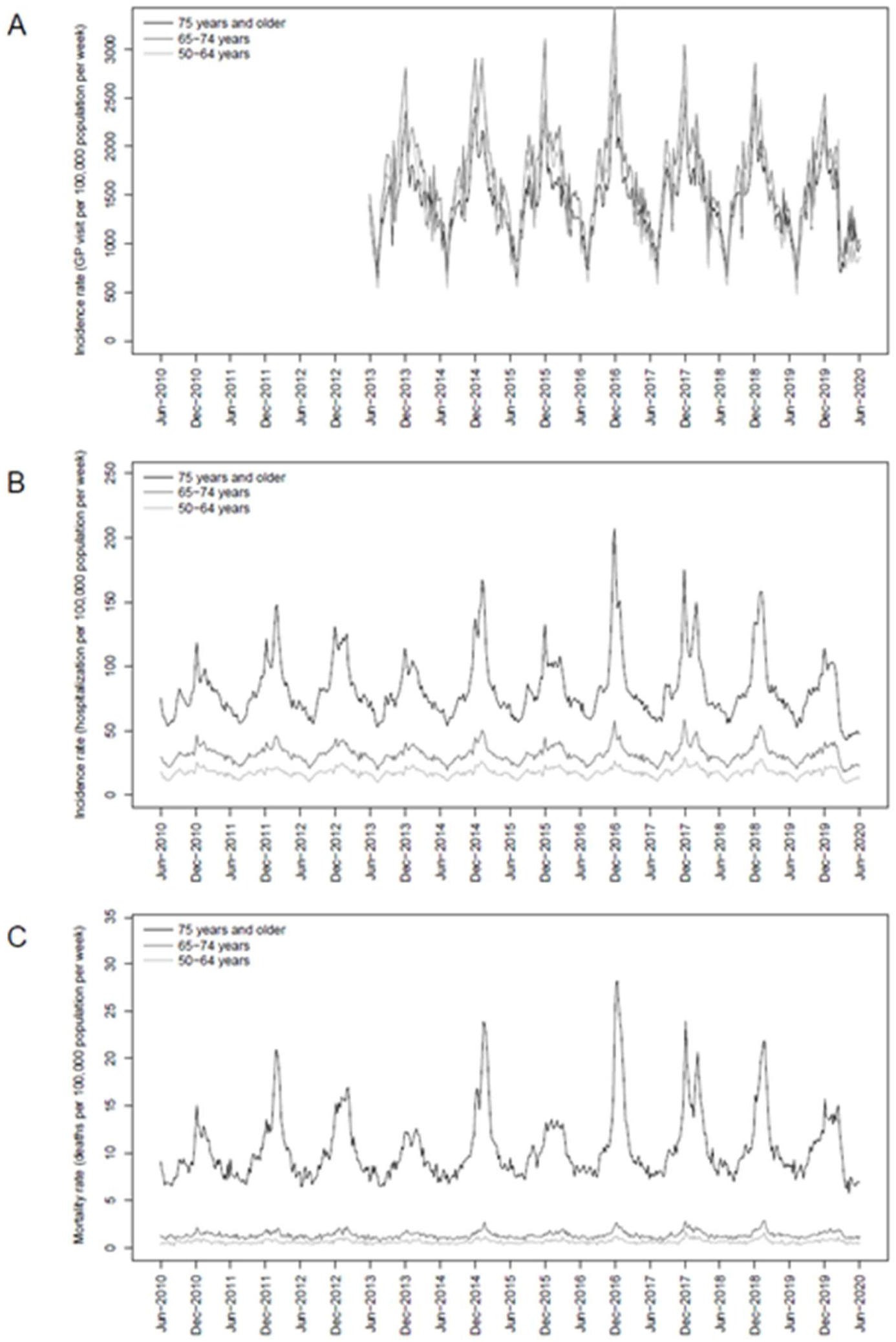
Weekly time-series of general practitioner visits, hospitalizations, and deaths by age group for respiratory causes (2010–2020). Evolution of the weekly number of general practitioner (GP) visits (A), hospitalizations (B), and deaths (C) per 100,000 population for respiratory causes (ICD-10 codes: J00–J99) as the primary diagnosis in France between June 2013 and June 2020 (A) or between June 2010 and June 2020 (B and C) for individuals aged 50–64, 65–74, or ≥75 years.

### Hospitalizations

Weekly numbers of hospitalizations for respiratory (ICD-10 codes: J00–J99), cardiorespiratory (ICD-10 codes: I00–I99, J00–J99), and narrowly-defined cardiorespiratory (ICD-10 codes: I21, I50, I63, I64, J00–J99) causes, stratified by age group, were extracted from the French national hospital database (Programme de Médicalisation des Systèmes d’Information [PMSI]) (Figure 1, Supplementary Figure 1, Supplementary Table 1). Hospitalizations were identified using the primary diagnosis code (i.e., the code used to describe the main reason for admission) alone and primary and secondary diagnoses codes (i.e., the codes used to report complications during hospitalization, the results of laboratory tests, or any comorbidities). For the characterisation and economic burden estimation of RSV hospitalizations, we included adults aged ≥50 years with at least one primary or secondary RSV- (i.e., J121, J205, J210, B974), or influenza-related (i.e., J09, J10, J11) ICD-10 code (Supplementary Table 1).

### Deaths

Weekly numbers of deaths from respiratory (ICD-10 codes: J00–J99) or cardiorespiratory (ICD-10 codes: I00–I99, J00–J99) causes were extracted from the National Epidemiology Centre of Medical Causes of Death (CépiDc) database, which collects death certificates for the entire French population (19).

### Study period

Data on the weekly numbers of hospitalizations and deaths were extracted from the PMSI and the CépiDc databases from the first week of July 2010 to the last week of June 2020 (spanning 10 epidemic seasons). Data on the weekly number of GP visits were collected from the first week of July 2013 to the last week of June 2020 from the EMR database due to data access restrictions. For modelling, the last week of February 2020 was set as the data cut-off date to exclude the COVID-19 pandemic period.

### Indicators of respiratory syncytial virus and influenza activity

The weekly numbers of hospitalizations due to RSV (ICD-10 codes: J20.5, J21.0, J21.9) among children aged <2 years and due to influenza (ICD-10 codes: J09, J10, J11) among individuals aged ≥65 years were extracted from the PMSI database as indicators of RSV and influenza circulation, respectively (Figure 2 and Supplementary Table 1).

**Figure 2.**
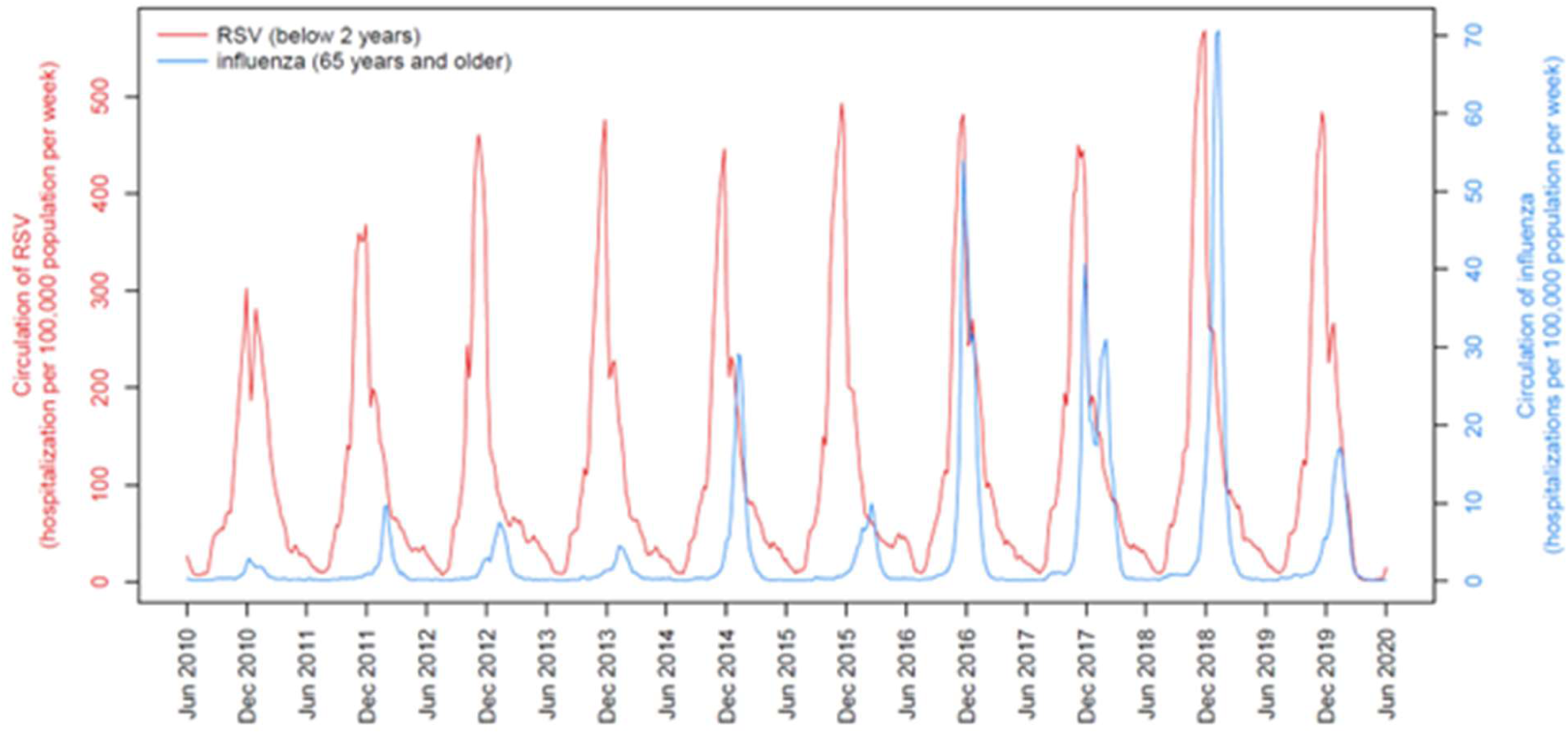
Weekly time-series of respiratory syncytial virus and influenza circulation (2010–2020). Evolution of the weekly number of hospitalizations per 100,000 population due to respiratory syncytial virus (RSV) infection among children aged ≤2 years (ICD-10 codes: J205, J210, J219) or influenza infection among adults aged ≥65 years (ICD-10 codes: J09, J10, J11) in France between June 2010 and June 2020.

Further information on the model, incidence estimation, and data sourcing can be found in the Supplementary Materials.

### Estimation of hospitalization costs

Average annual hospitalization costs were calculated from public health expenditure perspective, using the PMSI database. Economic burdens attributable to RSV and influenza were estimated by multiplying the mean hospital cost per RSV- or influenza-associated hospital stay by the estimated incidence of hospitalization. As RSV hospitalization costs varied significantly across seasons due to testing and coding changes, the final analysis used data from the last two RSV seasons (2017–2019), considered more representative of actual costs.

### Statistical analysis

All statistical analyses were performed using SAS software, version 9.4 (SAS Institute Inc., Cary, NC, USA) and R, version 4.2 .0 (The R Foundation, Vienna, Austria). Cyclic Poisson regression models were used to estimate the incidence rates of GP visits, hospitalizations, and deaths attributable to RSV and influenza infection. Weekly numbers of GP visits, hospitalizations, and deaths were modelled using a log link function to account for the baseline incidence of each outcome, time trends, seasonal terms, and independent indicators of RSV and influenza activity. Age- and cause-specific hospitalization data were interpreted using RSV/influenza circulation activity, time trends, and seasonal term.

## RESULTS

### Incidence of general practitioner visits

Over 2013–2020, the estimated average annual incidence rate of GP visits attributable to RSV in adults aged ≥65 years was 4,829/100,000 population for respiratory causes and 5,448/100,000 for cardiorespiratory causes (Table 1). This corresponded to 573,998 and 647,619 GP visits, representing 5.84% and 5.50% of total annual GP visits for this age group for respiratory and cardiorespiratory causes, respectively (Supplementary Table 3). These figures rose to 10.81% and 10.44% when restricting the timeframe to November to March each year (Northern Hemisphere winter). The incidence rate for respiratory causes varied from 2,736/100,000 among adults aged 50–64 years to 5,396/100,000 among adults aged 65–75 years (Table 2, Supplementary Figure 3). The mean annual incidence of influenza-attributable GP visits was lower among adults aged ≥65 years (2,469/100,000 for respiratory causes and 2,488/100,000 for cardiorespiratory causes).

**Table 1.**
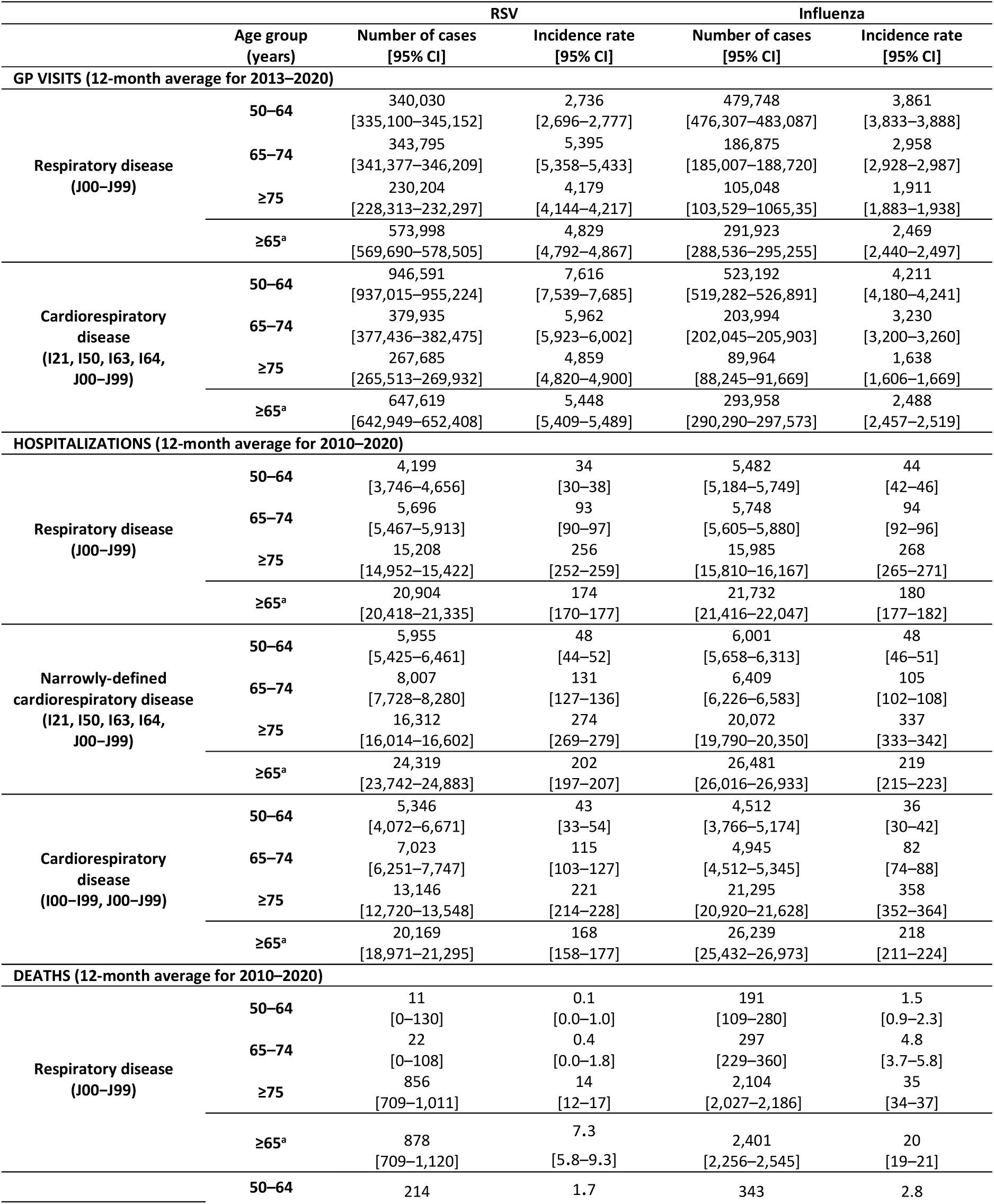
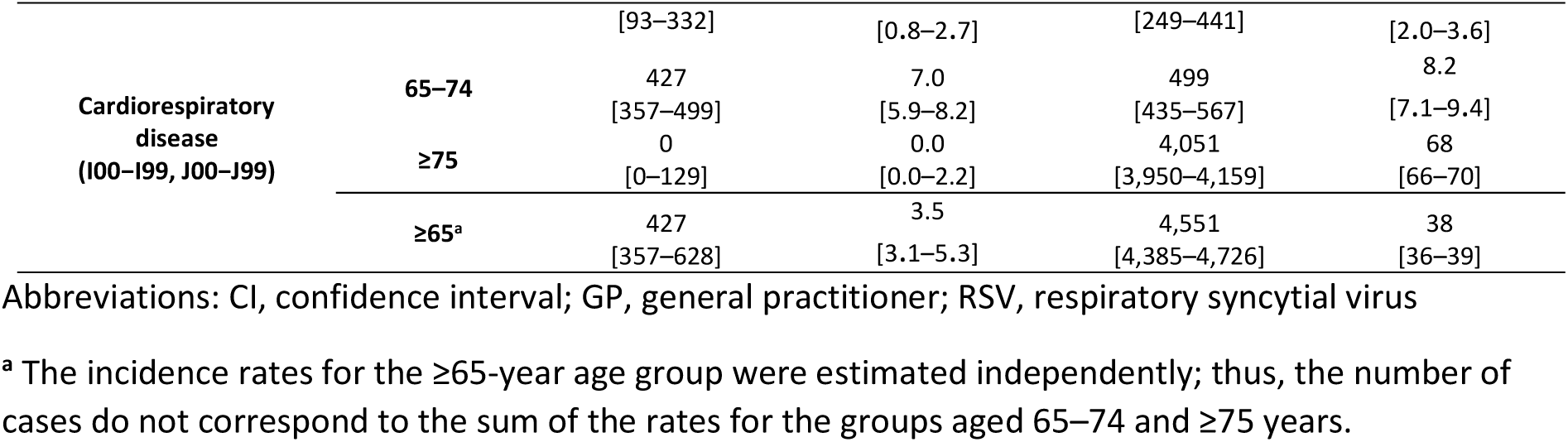
Mean estimated incidence of general practitioner visits, hospitalizations, and deaths attributable to respiratory syncytial virus and influenza infections, using respiratory or cardiorespiratory primary diagnosis ICD-10 codes.

**Table 2.** Average estimated incidence of hospitalizations attributable to respiratory syncytial virus and influenza infections, using respiratory or cardiorespiratory primary and secondary diagnoses ICD-10 codes.

|  |  | RSV |  | Influenza |  |
| --- | --- | --- | --- | --- | --- |
|  | Age group<br>(years) | Number of cases<br>[95% CI] | Incidence rate<br>[95% CI] | Number of cases<br>[95% CI] | Incidence rate<br>[95% CI] |
| HOSPITALIZATIONS (12-MONTH average for 2010-2020) |  |  |  |  |  |
| Respiratory disease<br>(J00-J99) | 50-64 | 700<br>[183-1,212] | 5.6<br>[1.5-9.8] | 5,662<br>[5,157-6,119] | 46<br>[42-49] |
|  | 65-74 | 9,255<br>[8,770-9,748] | 152<br>[144-160] | 6,489<br>[6,178-6,761] | 107<br>[101-111] |
|  | ≥75 | 17,256<br>[16,924-17,562] | 290<br>[285-296] | 22,365<br>[22,060-22,653] | 376<br>[371-381] |
|  | ≥65<sup>a</sup> | 26,511<br>[25,694-27,309] | 220<br>[214-227] | 28,855<br>[28,238-29,414] | 239<br>[234-244] |
| Narrowly-defined<br>cardiorespiratory disease<br>(I21, I50, I63, I64, J00-J99) | 50-64 | 8,437<br>[7,608-9,267] | 68<br>[61-75] | 6,290<br>[5,738-6,766] | 51<br>[46-55] |
|  | 65-74 | 12,015<br>[11,491-12,543] | 197<br>[189-206] | 7,586<br>[7,253-7,883] | 125<br>[119-130] |
|  | ≥75 | 21,926<br>[21,519-22,298] | 369<br>[362-375] | 29,045<br>[28,693-29,370] | 488<br>[482-494] |
|  | ≥65<sup>a</sup> | 33,941<br>[33,010-34,841] | 282<br>[275-290] | 36,631<br>[35,946-37,252] | 304<br>[298-309] |
| Cardiorespiratory disease<br>(I00-I99, J00-J99) | 50-64 | 6,956<br>[4,850-9,119] | 56<br>[39-74] | 3,059<br>[2,227-4,033] | 25<br>[18-33] |
|  | 65-74 | 12,713<br>[11,327-14,183] | 210<br>[187-234] | 3,283<br>[2,637-3,843] | 56<br>[45-65] |
|  | ≥75 | 0<br>[0-0] | 0.0<br>[0.0-0.0] | 18,139<br>[17,519-18,669] | 305<br>[295-314] |
|  | ≥65<sup>a</sup> | 12,713<br>[11,327-14,183] | 106<br>[94-118] | 21,422<br>[20,156-22,511] | 178<br>[168-188] |

Abbreviations: CI, confidence interval; RSV, respiratory syncytial virus
<sup>a</sup> The incidence rates for the ≥65-year age group were estimated independently; thus, the number of cases did not correspond to the sum of the rates for the groups aged 65-74 and ≥75 years.

### Incidence of hospitalizations

Between 2010 and 2020, in adults aged ≥65 years, the mean annual hospitalization incidence rate attributable to RSV as primary diagnosis code was 174/100,000 for respiratory causes, 202/100,000 for narrowly-defined cardiorespiratory causes, and 168/100,000 for cardiorespiratory causes (Table 1). This corresponded to 6.00%, 3.26%, and 1.56% of annual hospitalizations for respiratory, narrowly-defined cardiorespiratory, and all cardiorespiratory causes, respectively, and 11.37%, 6.57%, and 3.23% when restricting the analysis to November-March (Supplementary Table 3). Hospitalization rates due to respiratory causes increased with age (50–64 years: 34/100,000; 65–74 years: 93/100,000; ≥75 years: 256/100,000) (Table 1, Supplementary Figure 3). Average annual hospitalizations for narrowly-defined cardiorespiratory disease for adults aged ≥65 years was 24,319 [23,742–24,883] (Table 1). Mean annual hospitalization incidence rates for influenza and RSV were comparable (Table 1).

When including secondary diagnosis codes, we found that 17.7% of patients hospitalized with RSV as a secondary diagnosis had no respiratory or cardiac codes as primary diagnosis (Supplementary Table 4) and were not captured in the primary code analysis. When both primary and secondary diagnosis codes were considered, the annual mean incidence rate of RSV-attributable hospitalization in adults aged ≥65 years was 220/100,000 for respiratory causes and 282/100,000 for narrowly-defined cardiorespiratory causes (Table 2), representing a 26% and 40% increase, respectively, compared to primary diagnosis codes alone. **Mortality**

Over 2010–2020, the estimated mean annual mortality rate attributable to RSV in adults aged ≥65 years was 7.3/100,000 for respiratory causes and 3.5/100,000 for cardiorespiratory causes, corresponding to 878 and 427 deaths per year (Table 1). The mean annual mortality rate due to RSV-associated respiratory causes increased with age (50–64 years: 0.1 deaths per 100,000; 65-74 years: 0.4/100,000; ≥75 years: 14/100,000) (Table 1, Supplementary Figure 3). Annual mortality rates were higher for cardiorespiratory causes than for respiratory causes in the 50–64 and 65–74 age groups (Table 1). In individuals aged ≥75 years, RSV mortality estimates for cardiorespiratory causes were not generated due to poor model fit, impacting the estimates among adults ≥65 years. Overall, 2.47% of the annual, and 4.52% of seasonal deaths for respiratory causes were attributable to RSV (Supplementary Table 3). The estimated average annual mortality attributable to influenza in adults aged ≥65 years was higher than that attributable to RSV (20/100,000 for respiratory causes and 38/100,000 for cardiorespiratory causes) (Table 1).

### Hospitalization characteristics

Directly examining RSV and influenza hospitalizations from 2015 to 2019 diagnosed by standard-of-care testing, we identified 10,048 RSV-related and 108,657 influenza-related hospitalizations among adults aged ≥50 years (Table 3). For adults aged ≥65 years, there were 8,183 and 87,867 hospitalizations coded for RSV and influenza respectively (Table 3). Annual RSV hospitalizations for adults aged ≥65 years were underestimated by a factor of around 12 compared with the modelling approach. We found that 60.8% of RSV hospitalizations and 60.2% of influenza hospitalizations involved patients aged ≥75 years (Table 3).

**Table 3.** Characterization of hospitalizations with respiratory syncytial virus diagnoses (ICD-10 codes: J12.1, J20.5, J21.0, B974) or influenza diagnoses (ICD-10 codes: J09, J10, J11) infection, by age group (2015–2019)

|  | RSV |  |  |  | Influenza |  |  |  |
| --- | --- | --- | --- | --- | --- | --- | --- | --- |
|  | Aged 50–64 years<br>(N=1,865) | Aged 65–74 years<br>(N=2,071) | Aged ≥75 years<br>(N=6,112) | Aged ≥65 years<br>(N=8,183) | Aged 50–64 years<br>(N=20,790) | Aged 65–74 years<br>(N=22,381) | Aged ≥75 years<br>(N=65,486) | Aged ≥65 years<br>(N=87,867) |
| **Duration of hospitalization (days)** |  |  |  |  |  |  |  |  |
| Mean (SD) | 12.3 (15.2) | 12.5 (14.2) | 12.0 (10.4) | 12.1 (11.5) | 10.0 (14.8) | 10.6 (15.2) | 10.8 (10.2) | 10.7 (11.7) |
| Median (IQR) | 8.0 (4.0–14.0) | 8.0 (5.0–15.0) | 9.0 (6.0–15.0) | 9.0 (5.0–15.0) | 6.0 (2.0–11.0) | 7.0 (3.0–13.0) | 8.0 (4.0–14.0) | 8.0 (4.0–14.0) |
| Min–Max | 0–192 | 0–145 | 0–124 | 0–145 | 0–305 | 0–898 | 0–186 | 0–898 |
| **Disease severity, N (%)** |  |  |  |  |  |  |  |  |
| At least one stay in intensive care, continuous care, or resuscitation | 696 (37.3) | 742 (35.8) | 1,162 (19.0) | 1,904 (23.3) | 6,223 (29.9) | 5,945 (26.6) | 8,789 (13.4) | 14,734 (16.8) |
| In-hospital deaths | 93 (5.0) | 130 (6.3) | 558 (9.1) | 688 (8.4) | 933 (4.5) | 1,332 (6.0) | 5,572 (8.5) | 6,904 (7.9) |
| **3-month readmission, N (%)** |  |  |  |  |  |  |  |  |
| All causes | 931 (52.5) | 968 (49.9) | 2,140 (38.5) | 3,108 (41.5) | 7,061 (35.6) | 8,123 (38.6) | 19,329 (32.3) | 27,452 (33.9) |
| Cardiorespiratory causes | 291 (16.4) | 316 (16.3) | 862 (15.5) | 1,178 (15.7) | 1,488 (7.5) | 1,983 (9.4) | 5,976 (10.0) | 7,959 (9.8) |
| Respiratory causes | 261 (14.7) | 263 (13.5) | 588 (10.6) | 851 (11.4) | 1,247 (6.3) | 1,514 (7.2) | 3,794 (6.3) | 5,308 (6.6) |

Abbreviations: SD; standard deviation; IQR, interquartile range; RSV, respiratory syncytial virus

In patients aged ≥65 years, the average length of hospital stay was 12.1 days and 10,7 days for RSV and influenza hospitalizations respectively. The proportion of patients hospitalized for RSV requiring intensive care, continuous care, or resuscitation units decreased with age (50–64 years: 37.3%; 65–74 years: 35.8%; ≥75 years: 19.0%) (Table 3). A similar trend was observed for influenza hospitalizations with lower rates. In-hospital mortality increased with age for both RSV (50–64 years: 5.0%; ≥75 years: 9.1%), and influenza (50–64 years: 4.5%; ≥75 years: 8.5%).

### Economic burden of respiratory syncytial virus and influenza-attributable hospitalizations

Among patients aged ≥50 years, the average annual cost of hospitalization attributable to RSV infection was 130.9 millions € for hospitalizations with a respiratory cause and 43.9 millions € for hospitalizations with a cardiac cause, both as the main diagnosis (Table 4). Patients aged ≥65 years accounted respectively for 80.1% and 59,5% of the total costs. Among patients aged ≥50 years, the average annual cost of hospitalization attributable to influenza infection was 132.9 million € for hospitalizations with a respiratory cause and 31.4 million € for hospitalizations with a cardiac cause, both as the main diagnosis. Hospitalization cost estimates for influenza were similar to those for RSV (Table 4)

**Table 4.** Economic burden of respiratory syncytial virus and influenza hospitalizations, by age group.

|  | Age group |  |  |  |  |
| --- | --- | --- | --- | --- | --- |
|  | 50–64 years | 65–74 years | ≥75 years | ≥50 years (All) | ≥65 years |
| **RSV (2017–2019)** |  |  |  |  |  |
| **Average cost of RSV hospitalization with a primary respiratory diagnosis (€)** |  |  |  |  |  |
| Mean (SD) | 6,167 (6,662) | 5,908 (6,354) | 4,656 (4,197) | 5,199 (5,274) | 4,988 (4,894) |
| Median (IQR) | 4,261<br>(2,692–6,710) | 3,971<br>(3,041–6,458) | 3,374<br>(3,045–4,777) | 3,763<br>(3,041–5,402) | 3,472<br>(3,041–4,948) |
| Min–Max | [311–63,890] | [567–66,839] | [271–63,775] | [271–66,839] | [271–66,839] |
| **Overall annual cost<sup>a</sup> attributable to RSV hospitalizations with a primary respiratory diagnosis (million €)** |  |  |  |  |  |
| Mean [95%CI] | 25.9<br>[23.1–28.7] | 33.7 [32.3–<br>34.9] | 70.8<br>[69.6–71.8] | 130.4<br>[125–135.5] | 104.5<br>[101.9–106.7] |
| **Average cost of an RSV hospitalization with a primary cardiac diagnosis (€)** |  |  |  |  |  |
| Mean (SD) | 10,119 (16,143) | 8,762 (9,248) | 5,324 (3,135) | 6,218 (6,634) | 5,866 (4,820) |
| Median (IQR) | 4,916<br>(3,516–8,786) | 5,695<br>(4,483–9,014) | 4,489<br>(3,312–6,515) | 4,502<br>(3,488–6,717) | 4,502<br>(3,365–6,717) |
| Min–Max | [838–107,847] | [666–54,220] | [531–39,183] | [531–107,847] | [5,31–54,220] |
| **Overall annual cost<sup>a</sup> attributable to RSV hospitalizations with a primary cardiac diagnosis<sup>b</sup> (million €),** |  |  |  |  |  |
| Mean [95%CI] | 17.8<br>[17–18.3] | 20.2 [19.8–<br>20.7] | 5.9<br>[5.7–6.3] | 43.9<br>[42.5–45.3] | 26.1<br>[25.5–27.0] |
| **INFLUENZA (2015–2019)** |  |  |  |  |  |
| **Cost of influenza hospitalization with a primary respiratory diagnosis (€)** |  |  |  |  |  |
| Mean (SD) | 5,624 (9,085) | 5,499 (7,675) | 4,413 (3,899) | 4,868 (6,112) | 4,691 (5,156) |
| Median (IQR) | 2,684<br>(15,68–5,050) | 4,326<br>(1,997–4,777) | 4,326<br>(2,694–4,396) | 4,326<br>(2,638–4,396) | 4,326<br>(2,638–4,396) |
| Min–Max | [212–192,592] | [164–142,932] | [212–121,665] | [164–192,592] | [164–142,932] |
| **Overall annual cost<sup>a</sup> attributable to influenza hospitalizations with primary respiratory diagnosis (million €)** |  |  |  |  |  |
| Mean [95%CI] | 30.8<br>[29.2–32.3] | 31.6 [30.8–<br>32.3] | 70.5<br>[69.8–71.3] | 132.9<br>[129.8–135.9] | 102.1 [100.6–103.64] |
| **Cost of influenza hospitalization with a primary cardiac diagnosis (€)** |  |  |  |  |  |
| Mean (SD) | 8,099 (11,117) | 7,594 (9,632) | 5,437 (3,948) | 6,163 (6,658) | 5,890 (5,706) |
| Median (IQR) | 4,677<br>(2,975–8,359) | 4,681<br>(3,294–8,139) | 4,502<br>(3,468–6,617) | 4,502<br>(3,312–6,732) | 4,502<br>(3,426–6,717) |
| Min–Max | [281–145,055] | [281–173,403] | [276–67,771] | [276–173,403] | [276–173,403] |
| **Overall annual cost<sup>a</sup> attributable to influenza hospitalizations with primary cardiac diagnosis<sup>b</sup> (million €)** |  |  |  |  |  |
| Mean [95%CI] | 4.2<br>[3.8–4.6] | 5.0<br>[4.7–5.3] | 22.2<br>[21.6–22.7] | 31.4<br>[30.1–32.6] | 27.2 [26.3–28.0] |

Abbreviations: CI, confidence interval; IQR, interquartile range; SD, standard deviation; RSV, respiratory syncytial virus
<sup>a</sup> Costs were estimated using the average hospitalization cost for patients aged ≥50 years with at least one RSV or influenza hospitalization. These per patient costs were applied to the number of excess cases estimated using the cyclic Poisson regression models for the 2010–2020 period (Table 2).
<sup>b</sup> The number of cardiorespiratory excess cases (ICD-10 codes: I00–I99, J00–J99) minus the number of respiratory excess cases (ICD-10 codes: J00–J99) estimated using cyclic Poisson regression models (Table 2).

## DISCUSSION

We estimated the incidence of RSV-attributable GP visits, hospitalizations and mortality, among adults aged ≥65 years in France. Previous hospitalization estimates based on RSV-specific ICD-10 codes have underestimated RSV burden due to limited testing, low diagnostic accuracy, and misclassification (8,20). Using a model-based approach, we estimated ∼25,000 RSV hospitalizations annually in this population— about 13 times higher than what ICD-10 coding alone would suggest (∼2,000 cases/year).

RSV hospitalization incidence among adults ≥65 years in this study ranged from 174 to 282/100,000. Higher rates were reported in the U.K., Spain, and Germany (234–787/100,000) (4,5,11). A pooled analysis of U.S. studies reported an adjusted RSV hospitalization incidence rate for this population of 282/100,000 for prospective studies and 235.7/100,000 for time-series analyses (21) after adjustment for diagnostic testing-based under ascertainment. A another meta-analysis reported a lower pooled rate of 150/100,000 among individuals aged ≥65 years (3), though this result was influenced by the healthy volunteer effect of clinical trials and methodological variations (primary and secondary diagnoses, inclusion of respiratory and cardiorespiratory causes of hospitalization). In our analyses, we examined the impact of parameters on the model, and the incidence rates. Standardizing methodologies across countries could improve comparability.

Influenza hospitalization rates among adults ≥65 years ranged from 180 to 304/100,000. Previous studies estimated influenza hospitalization incidence in France using the PMSI), with rates ranging from 36/100,000 (60-79 years) to 134/100,000 (80 years and over) ove*r* 2012-2017 (22). Lemaitre et al. reported broader seasonal variability in influenza hospitalizations for respiratory causes in adults ≥65 years over 2010-2018, with annual rates ranging from 26 to 308 /100,000 depending to years, using a similar model approach (23).

Our study also provides one of the few estimates of RSV-related GP visits. The average annual incidence rates ranged from 2,000–8,000/100,000, higher in the 50–64-year age group than in the ≥65-year age group. The incidence rate of RSV-attributable GP visits was twice higher than that attributable to influenza. This higher rate of RSV in primary care should be considered when evaluating the strains on primary care services. These observations do not take into account the higher vaccination rate for influenza among the older population.

Mortality rates increased with age ranging from 0.1–14/100,000. This highlights the older adults’ vulnerability to severe disease from respiratory viruses such as RSV. These mortality rates were lower than the 37.9-44.3/100,000 observed by Haeberer *et al.* for adults aged ≥60 years (5) but similar to those reported by Hansen *et al.* for excess mortality in adults aged ≥65 years for RSV (14.7/100,000) and influenza (20.5/100,000) (24).

Comparing RSV severity to influenza severity, we observed much lower mortality for RSV than influenza mortality. Although hospitalization rates for RSV and influenza were similar, RSV hospitalizations appeared more severe when analysing hospitalization stay characteristics. Patients with an RSV code were hospitalized for longer and were more likely to be admitted to intensive care, continuous care, or resuscitation units than those who did not have a diagnosis of RSV. These results may reflect the lack of routine testing for RSV in France, where routine testing is only performed in intensive care units, leading to the systematic capture of only the most severe RSV cases.

Both RSV and influenza can cause respiratory and cardiac events (25), and our incidence rates for GP visits and hospitalizations were ∼20% higher when we evaluated cardiorespiratory rather than respiratory symptoms alone. However, broad ICD-10 cardiac codes (I00–99) previously used to generate the time-series proved unreliable during modelling. For example, the estimated number of cardiorespiratory cases was lower than the number of respiratory cases. To increase model performance, we restricted our search to narrowly-defined cardiorespiratory conditions (identified using a limited set of diagnostic codes), which are known to be linked to RSV and influenza infections. Although these narrow definition cardiorespiratory codes were successfully used for the estimation of RSV-attributable GP visits and hospitalizations, we could not obtain adequate data to inform the mortality rate analysis. Nevertheless, these findings demonstrate how model accuracy can be improved by restricting the definition of diagnostic codes to reduce noise in the model.

Secondary diagnosis codes also merit consideration. While primary diagnosis codes reflect hospital admission reasons, secondary diagnosis codes are used to better characterize hospital stays. Our principal analysis used only primary codes, as secondary codes aren’t recorded for GP visits and death. However, using both primary and secondary codes for hospitalizations increased our estimates by 26-40%. A recent systematic literature review of ICD code validity studies found that use of only primary ICD codes miss a number of hospitalizations for pneumonia and lower respiratory tract infection events (26). Analysis of hospital stays including an RSV ICD-10 code in any position revealed that most codes primary ICD-10 codes were related to respiratory and cardiac manifestations. This finding reinforces the value of including cardiac codes in the analysis. However, the top 10 primary codes outside the respiratory and cardiac categories were linked to pathologies unrelated to RSV infection (e.g. oncology). Thus, considering both primary and secondary ICD-10 codes not only capture RSV infection leading to hospitalisation but also RSV infection in hospitalized patients. Further research is needed to better understand the impact of RSV infection during hospitalization.

The estimated costs for respiratory hospitalizations attributable to RSV or influenza among patients aged ≥65 years were comparable (Supplementary Table 5). The mean total costs for cardiac hospitalizations attributable to RSV or influenza were 124 million € and 130 million € among patients aged >65 years, respectively, which aligns with the findings of recent influenza study (23).

Influenza infections displayed more seasonal variability than RSV, also observed in a U.S. study evaluating excess influenza and RSV mortality from 1999-2018 (24). This likely reflects the circulation of distinct influenza subtypes (H1N1, H3N2, B), which varying severity by age and across season. H3N2 strains typically cause more severe disease in older adults, while B and H1N1 strains affect younger individuals (24,27). Other factors, like antigenic shift or drift, can create new strains capable of evading preexisting immunity, while virus-vaccine mismatches can reduce vaccine effectiveness(28).

Model-based studies have inherent limitations. The use of different virus circulation indicators can significantly affect estimates. The estimates were modelled using bronchiolitis hospitalizations in children aged <2 years and influenza hospitalizations in adults aged ≥65 years. We retained all relevant codes from a study showing that RSV bronchiolitis was coded via J205, J210, and J219 (29). We carried out sensitivity analyses (data not shown) using proxies from emergency room visits for bronchiolitis in children aged <2 years for RSV, and influenza-like illness for influenza from the sentinel GP network. Although influenza estimates remained unchanged, RSV data were significantly higher. Therefore, we adopted a conservative approach, relying on proxies derived solely from hospital data. We also investigated the impact of lags on the estimations (see supplementary materials). Our model could not capture the burden of other seasonal virus like parainfluenza due to the unavailability of robust data, nor could it assess RSV related emergency visits. Further research on these two topics would be beneficial.

## CONCLUSIONS

This study highlights the significant burden of RSV infection in adults ≥65 years in France. The incidence rate and economic burden of hospitalization were similar to the current burden of influenza. This work underscores the need for model-based studies to better estimate and compare the burdens of respiratory diseases and generate real-world evidence to support prevention campaign implementation. Given the substantial clinical and economic burden of RSV infection, measures, such as vaccination campaigns, should be taken to prevent its spread.

## Data Availability

The national datasets used in this study can be requested from the appropriate French authorities, the model datasets used for analysis are not publicly available.

## Acknowledgements

The authors thank Dan Weinberger, Associate Professor in Epidemiology of Microbial Diseases at Yale School of Public Health for advising on the selection of the methodology used in this study. The authors are grateful to the Direction de la Stratégie des Études et des Statistiques (DSES), Département Accès, Traitement et Analyse de la Donnée (DATAD), and Cellule de la CNAM en Charge de l’accompagnement des Demandes D’extraction (DEMEX) teams, especially Julien Brande and Marjorie Boussac, at the Caisse Nationale de l’Assurance Maladie (CNAM) for their assistance with data extraction.

## Author contributions

CN and ML contributed to the study conception and design, as well as the interpretation of data and critical review of the manuscript. VB, CBF, EL, and LW contributed to the methodology and data analysis. SF, EB, HL, PV, JSC, LW, PL, CL, and EB contributed to the study design, and interpretation of data. All authors reviewed and approved the final manuscript ahead of submission.

## Manuscript preparation

HORIANA and Anya Lissana provided assistance in preparing and editing the manuscript.

## Data availability

The datasets used in this study are not publicly available.

## REFERENCES

1. Falsey AR, Hennessey PA, Formica MA, Cox C, Walsh EE. Respiratory syncytial virus infection in elderly and high-risk adults. N Engl J Med. 2005 Apr 28;352(17):1749–59.

2. Njue A, Nuabor W, Lyall M, Margulis A, Mauskopf J, Curcio D, et al. Systematic Literature Review of Risk Factors for Poor Outcomes Among Adults With Respiratory Syncytial Virus Infection in High-Income Countries. Open Forum Infect Dis. 2023 Nov;10(11):ofad513.

3. Savic M, Penders Y, Shi T, Branche A, Pirçon JY. Respiratory syncytial virus disease burden in adults aged 60 years and older in high-income countries: A systematic literature review and meta-analysis. Influenza Other Respir Viruses. 2023;17(1):e13031.

4. Polkowska-Kramek A, Begier E, Bruyndonckx R, Liang C, Beese C, Brestrich G, et al. Estimated Incidence of Hospitalizations and Deaths Attributable to Respiratory Syncytial Virus Infections Among Adults in Germany Between 2015 and 2019. Infect Dis Ther. 2024 Apr;13(4):845–60.

5. Haeberer M, Bruyndonckx R, Polkowska-Kramek A, Torres A, Liang C, Nuttens C, et al. Estimated Respiratory Syncytial Virus-Related Hospitalizations and Deaths Among Children and Adults in Spain, 2016–2019. Infect Ther. 2024;13:463–80.

6. Li Y, Kulkarni D, Begier E, Wahi-Singh P, Wahi-Singh B, Gessner B, et al. Adjusting for Case Under-Ascertainment in Estimating RSV Hospitalisation Burden of Older Adults in High-Income Countries: a Systematic Review and Modelling Study. Infect Dis Ther. 2023 Mar 20;1–13.

7. Bruyndonckx R, Polkowska-Kramek A, Liang C, Nuttens C, Tran TMP, Gessner BD, et al. Estimation of Symptomatic Respiratory Syncytial Virus Infection Incidence in Adults in Multiple Countries: A Time-Series Model-Based Analysis Protocol. Infect Dis Ther. 2024 Apr;13(4):953–63.

8. Loubet P, Fernandes J, de Pouvourville G, Sosnowiez K, Elong A, Guilmet C, et al. Respiratory syncytial virus-related hospital stays in adults in France from 2012 to 2021: A national hospital database study. J Clin Virol. 2024 Apr 1;171:105635.

9. Binder W, Thorsen J, Borczuk P. RSV in adult ED patients: Do emergency providers consider RSV as an admission diagnosis? Am J Emerg Med. 2017 Aug;35(8):1162–5.

10. Chartrand C, Tremblay N, Renaud C, Papenburg J. Diagnostic Accuracy of Rapid Antigen Detection Tests for Respiratory Syncytial Virus Infection: Systematic Review and Meta-analysis. J Clin Microbiol. 2015 Dec;53(12):3738–49.

11. Fleming DM, Taylor RJ, Lustig RL, Schuck-Paim C, Haguinet F, Webb DJ, et al. Modelling estimates of the burden of Respiratory Syncytial virus infection in adults and the elderly in the United Kingdom. BMC Infect Dis. 2015 Oct 23;15:443.

12. Onwuchekwa C, Moreo LM, Menon S, Machado B, Curcio D, Kalina W, et al. Under-ascertainment of Respiratory Syncytial Virus infection in adults due to diagnostic testing limitations: A systematic literature review and meta-analysis. J Infect Dis. 2023 Jul 14;228(2):173–84.

13. Ramirez J, Carrico R, Wilde A, Junkins A, Furmanek S, Chandler T, et al. Diagnosis of Respiratory Syncytial Virus in Adults Substantially Increases When Adding Sputum, Saliva, and Serology Testing to Nasopharyngeal Swab RT-PCR. Infect Dis Ther. 2023 Jun 1;12(6):1593–603.

14. Rozenbaum MH, Begier E, Kurosky SK, Whelan J, Bem D, Pouwels KB, et al. Incidence of Respiratory Syncytial Virus Infection in Older Adults: Limitations of Current Data. Infect Dis Ther. 2023 Jun;12(6):1487–504.

15. Rozenbaum MH, Judy J, Tran D, Yacisin K, Kurosky SK, Begier E. Low Levels of RSV Testing Among Adults Hospitalized for Lower Respiratory Tract Infection in the United States. Infect Dis Ther. 2023 Feb;12(2):677–85.

16. Chow EJ, Rolfes MA, O’Halloran A, Anderson EJ, Bennett NM, Billing L, et al. Acute Cardiovascular Events Associated With Influenza in Hospitalized Adults. Ann Intern Med. 2020 Oct 20;173(8):605–13.

17. Kwong JC, Schwartz KL, Campitelli MA, Chung H, Crowcroft NS, Karnauchow T, et al. Acute Myocardial Infarction after Laboratory-Confirmed Influenza Infection. N Engl J Med. 2018 Jan 25;378(4):345–53.

18. Woodruff RC, Melgar M, Pham H, Sperling LS, Loustalot F, Kirley PD, et al. Acute Cardiac Events in Hospitalized Older Adults With Respiratory Syncytial Virus Infection. JAMA Intern Med. 2024 Jun 1;184(6):602–11.

19. Rey G. [Death certificate data in France: Production process and main types of analyses]. Rev Med Interne. 2016 Oct;37(10):685–93.

20. Cai W, Tolksdorf K, Hirve S, Schuler E, Zhang W, Haas W, et al. Evaluation of using ICD-10 code data for respiratory syncytial virus surveillance. Influenza Other Respir Viruses. 2020 Nov;14(6):630–7.

21. McLaughlin JM, Khan F, Begier E, Swerdlow DL, Jodar L, Falsey AR. Rates of Medically Attended RSV Among US Adults: A Systematic Review and Meta-analysis. Open Forum Infect Dis. 2022 Jul;9(7):ofac300.

22. Pivette M, Nicolay N, de Lauzun V, Hubert B. Characteristics of hospitalizations with an influenza diagnosis, France, 2012-2013 to 2016-2017 influenza seasons. Influenza Other Respir Viruses. 2020 May;14(3):340–8.

23. Lemaitre M, Fouad F, Carrat F, Crépey P, Gaillat J, Gavazzi G, et al. Estimating the burden of influenza-related and associated hospitalizations and deaths in France: An eight-season data study, 2010–2018. Influenza Other Respir Viruses. 2022 Jul;16(4):717–25.

24. Hansen CL, Chaves SS, Demont C, Viboud C. Mortality Associated With Influenza and Respiratory Syncytial Virus in the US, 1999-2018. JAMA Netw Open. 2022 Feb 28;5(2):e220527.

25. Ivey KS, Edwards KM, Talbot HK. Respiratory Syncytial Virus and Associations With Cardiovascular Disease in Adults. J Am Coll Cardiol. 2018 Apr 10;71(14):1574–83.

26. Hanquet G, Theilacker C, Vietri J, Sepúlveda-Pachón I, Menon S, Gessner B, et al. Best Practices for Identifying Hospitalized Lower Respiratory Tract Infections Using Administrative Data: A Systematic Literature Review of Validation Studies. Infect Dis Ther. 2024 Apr;13(4):921–40.

27. Carazo S, Guay CA, Skowronski DM, Amini R, Charest H, De Serres G, et al. Influenza hospitalization burden by subtype, age, comorbidity and vaccination status: 2012/13 to 2018/19 seasons, Quebec, Canada. Clin Infect Dis Off Publ Infect Dis Soc Am. 2023 Oct 11;ciad627.

28. Tricco AC, Chit A, Soobiah C, Hallett D, Meier G, Chen MH, et al. Comparing influenza vaccine efficacy against mismatched and matched strains: a systematic review and meta-analysis. BMC Med. 2013 Jun 25;11:153.

29. Demont C, Petrica N, Bardoulat I, Duret S, Watier L, Chosidow A, et al. Economic and disease burden of RSV-associated hospitalizations in young children in France, from 2010 through 2018. BMC Infect Dis. 2021 Aug 2;21:730.

