## Supplementary materials for "Estimation of GP visits, hospitalizations and deaths attributable to RSV and influenza and costs associated with hospitalizations in older adults in France, 2010-2020"

### **Data sources**

The cardiac codes used in this study were selected based on their associations with cardiac manifestations of respiratory syncytial virus (RSV) and influenza infection (1–3). The Electronic Medical Records (EMR) database collects electronic medical records from 1,200 general practitioners (GPs) in France (equivalent to ~2% of all French GPs) (4). The EMR database collects electronic medical records from 1,200 GPs in France (representing approximately 2% of all French GPs) (4). The panel of contributing GPs is selected to represent the primary care physician population based on three main criteria known to influence prescribing practices: age, sex, and geographical region. Hospitalizations were identified using the primary diagnosis code (i.e., the code used to describe main reason for hospitalization) and secondary diagnoses codes (i.e., the codes used to report complications during hospitalization, the results of laboratory tests, or any comorbidities). Mortality data were extracted from the National Epidemiology Center of Medical Causes of Death database, which collects death certificates for the entire French population (5).

### **Statistical analysis**

Age- and cause-specific hospitalization data were analyzed using a model that incorporated indicators of RSV/influenza activity, time trends, and the seasonal term variations. The model employed the following log link function:

$$\boldsymbol{log}\left( \boldsymbol{H}_{\boldsymbol{t}}^{\left( \boldsymbol{c,a} \right)} \right)\boldsymbol{=}\boldsymbol{\alpha}_{\boldsymbol{0}}\boldsymbol{+}\boldsymbol{\alpha}_{\boldsymbol{1}}\boldsymbol{t+}\boldsymbol{\alpha}_{\boldsymbol{2}}\boldsymbol{t}^{\boldsymbol{2}}\boldsymbol{+}\boldsymbol{\alpha}_{\boldsymbol{3}}\boldsymbol{t}^{\boldsymbol{3}}\boldsymbol{+}\sum_{\boldsymbol{i=1}}^{\boldsymbol{12}} \boldsymbol{[\beta}_{\boldsymbol{i}}\boldsymbol{cos}\left( \frac{\boldsymbol{it\pi}}{\boldsymbol{52,14}} \right)\boldsymbol{+}\boldsymbol{\gamma}_{\boldsymbol{i}}\boldsymbol{sin}\left( \frac{\boldsymbol{it\pi}}{\boldsymbol{52,14}} \right)\boldsymbol{]}\boldsymbol{+}\boldsymbol{\eta}\boldsymbol{lweek+}\sum_{\boldsymbol{i=1}}^{\boldsymbol{10}} \boldsymbol{\delta}_{\boldsymbol{i}}\boldsymbol{ILI}_{\boldsymbol{t-l}}\boldsymbol{+}\boldsymbol{\theta}\boldsymbol{RSV}_{\boldsymbol{t-l}}$$

where $\boldsymbol{H}_{\boldsymbol{t}}^{\left( \boldsymbol{c},\boldsymbol{a} \right)}$ is the weekly number of hospitalization or deaths from cause ‘c’ in age group ‘a’. Terms t, t², and t^3^ are the linear, quadratic, and cubic terms of the time trend, respectively. The terms cos and sin represent seasonality. A binary variable ***lweek***, which represents the last week of the year (as it includes two bank holidays: Christmas Day and New Year’s Day) was integrated into the model. $\boldsymbol{RSV}_{\boldsymbol{t}}$ and $\boldsymbol{ILI}_{\boldsymbol{t}}$ represent the weekly indicator of RSV and influenza activity, respectively. Parameters α, β, γ, δ, and θ were estimated using a likelihood method. Overdispersion was verified and alternative distributions (e.g., quasi-Poisson, binomial negative) were tested. The number of hospitalization and cause-specific hospitalization were included as raw data to take into consideration the impact of the indicators (i.e., RSV and influenza activity) on the weekly hospitalizations or deaths.

RSV and influenza circulation period (independent variable) were integrated into the model by using a variable for the entire period for RSV and by using an epidemic season for influenza. Several time period lags (4 lags for RSV and influenza) were tested to take into consideration the delay between RSV/influenza infection and the occurrence of a hospitalization/death event. A manual backward selection method was used to identify significant time trends and seasonal terms in each model. This method employed the Akaike Information Criterion (AIC) and correlations between the observed and predicted hospitalization values.Best model fit was obtained using data from GP visits and hospitalizations for respiratory causes; a 1-week lag was used for RSV and no lag was used for influenza for all age categories. Different time lags were observed between hospitalizations and deaths across age categories. For deaths, the best fit was achieved with a 2-week lag for RSV and 1-week lag for influenza for the 50–64-year and 65–74-year age groups. The mortality rate in patients aged ≥75 was estimated with a 3-week lag for RSV and a 2-week lag for influenza. The results are summarized in Supplementary Table 1.

The average incidence rates of RSV- and influenza-associated GP visits, hospitalizations, or deaths were estimated by calculating the difference between the predicted and baseline values for each outcome. These expected values were derived from Poisson models, whereby the RSV/influenza activity parameter was set to zero to establish the baseline. The total number of GP visits, hospitalizations, or deaths were calculated for each winter (November–March) for each season (12-months period) and overall period (average of all seasons); age-group-specific incidence rates were then determined. Confidence intervals of the estimated excesses were computed via bootstrap resampling of the model residuals (1,000 replicates). Both the annual and seasonal proportions of GP visits, hospitalizations, and deaths attributable to RSV or influenza were determined. Moreover, a descriptive analysis of patients hospitalized for RSV or influenza between 2015 and 2019 was performed.

A strong seasonal fluctuation in respiratory GP visits, hospitalizations, and deaths is typically observed. The increase in the number of respiratory hospitalizations during the cold season is characterized by two sequential peaks. During the study period, a 1–2-week lag was observed between the peaks of respiratory hospitalizations and deaths (Figure S1). Although the RSV epidemic periods were generally constant, with peaks observed across the same weeks each season, influenza epidemics were considerably more variable, with peaks occurring at different times within each cold season (Supplementary Figure 2).

We found significant estimation differences when lags were modified (from zero to three weeks, data not shown). We selected the lags using the best predicted model fit and determined the difference in circulation between children and adults by looking at the peak number of hospitalized bronchiolitis in children aged <2 years and the peak number of RSV coded hospitalization in adults in a group of hospital in Lyon, for two consecutive years (lag of one week observed).

### **Supplementary References**

1.	Chow EJ, Rolfes MA, O’Halloran A, Anderson EJ, Bennett NM, Billing L, et al. Acute Cardiovascular Events Associated With Influenza in Hospitalized Adults. Ann Intern Med. 2020 Oct 20;173(8):605–13.

2.	Kwong JC, Schwartz KL, Campitelli MA, Chung H, Crowcroft NS, Karnauchow T, et al. Acute Myocardial Infarction after Laboratory-Confirmed Influenza Infection. N Engl J Med. 2018 Jan 25;378(4):345–53.

3.	Woodruff RC, Melgar M, Pham H, Sperling LS, Loustalot F, Kirley PD, et al. Acute Cardiac Events in Hospitalized Older Adults With Respiratory Syncytial Virus Infection. JAMA Intern Med. 2024 Jun 1;184(6):602–11.

4.	Launay T, Souty C, Vilcu AM, Turbelin C, Blanchon T, Guerrisi C, et al. Common communicable diseases in the general population in France during the COVID-19 pandemic. PLoS ONE. 2021 Oct 11;16(10):e0258391.

5.	Rey G. [Death certificate data in France: Production process and main types of analyses]. Rev Med Interne. 2016 Oct;37(10):685–93.

### **Supplementary Figures and Tables**

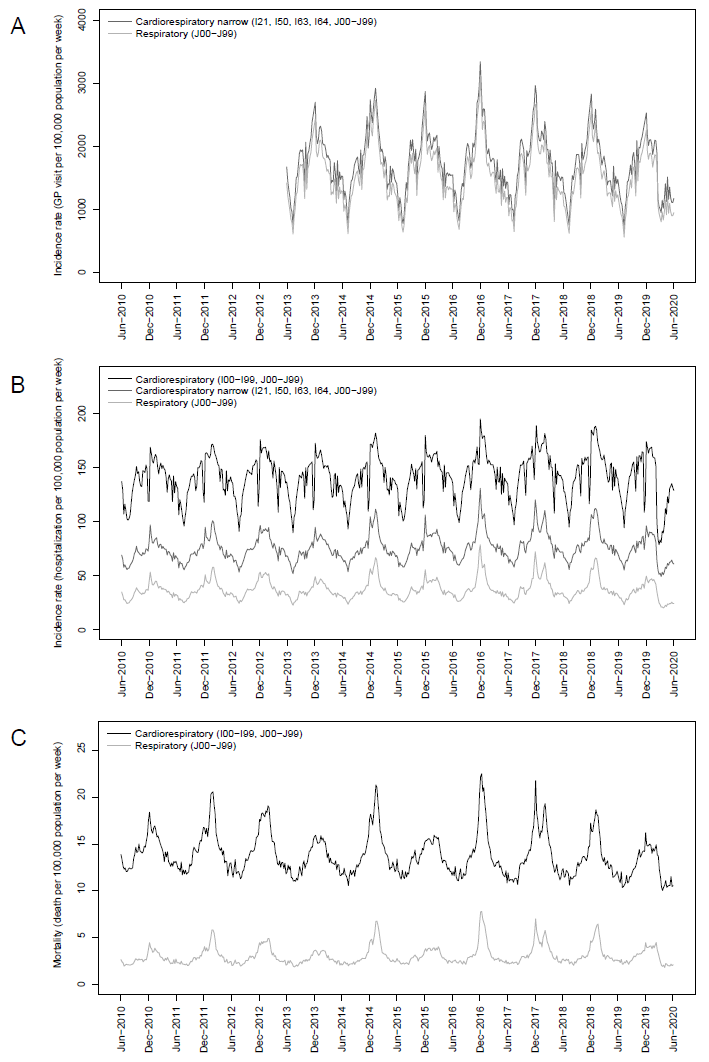

### **Supplementary Figure 1. Weekly time series of general practitioner visits, hospitalizations, and deaths for cardiorespiratory or respiratory** **ICD-10 codes, 2010–2020. Evolution of the weekly number of general practitioner (GP) visits (A), hospitalizations (B), and deaths (C) per 100,000 population for cardiorespiratory causes (ICD-10 codes: I00−I99, J00−J99), narrowly defined cardiorespiratory causes (ICD-10 codes: I21, I50, I63, I64, J00−J99), or respiratory causes (ICD-10 codes: J00–J99) as primary diagnosis in France between June 2013 to June 2020 (A) or June 2010 to June 2020 (B and C) among adults aged ≥50 years.**

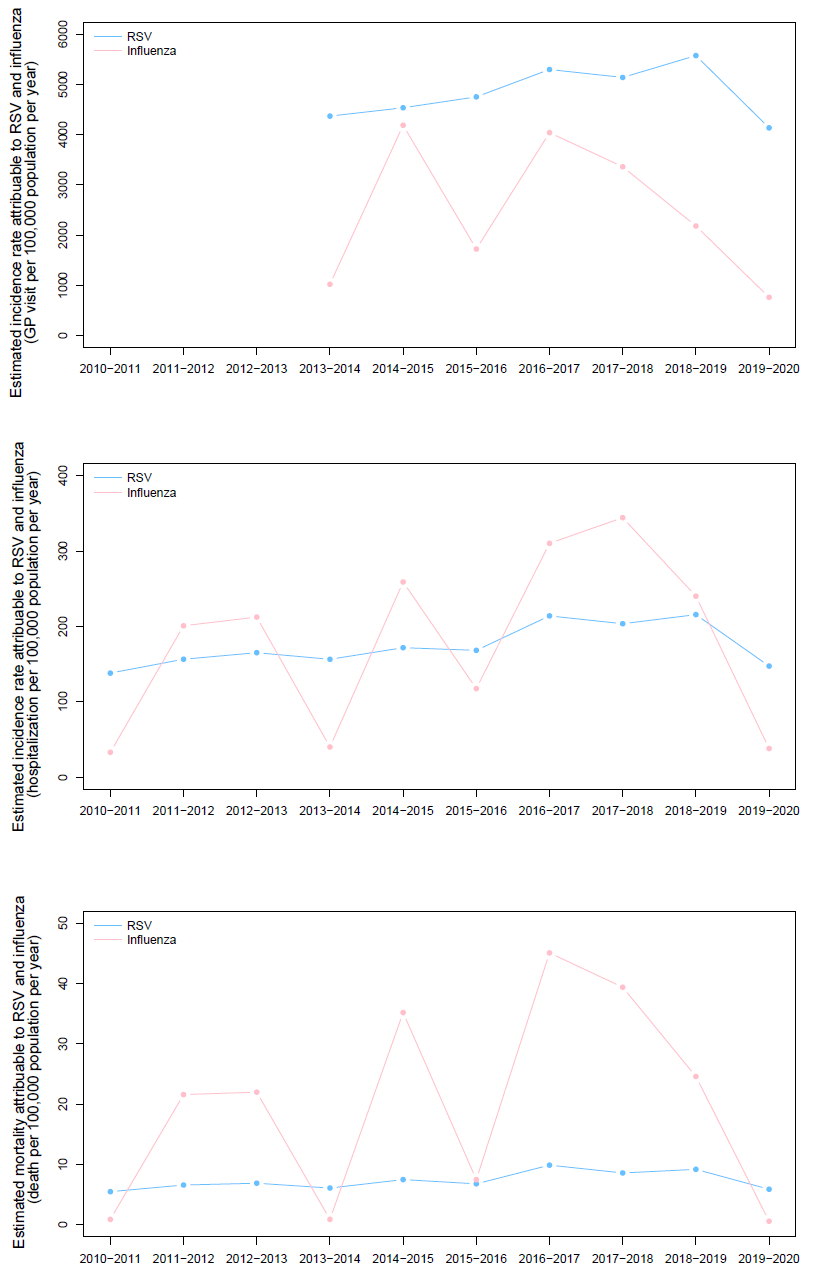

### **Supplementary Figure 2. Annual evolution of estimated excess general practitioner visits, hospitalizations, and deaths attributable to respiratory syncytial virus and influenza, 2010–2020. Estimated annual number of general practitioner (GP) visits (A), hospitalizations (B), and deaths (C) per 100,000 population for respiratory causes (ICD-10 codes: J00–J99 as the primary diagnosis) attributable to respiratory syncytial virus (RSV) or influenza in France from 2013–2014 to 2019–2020 (A) or from 2010–2011 to 2019–2020 (B and C) in patients aged ≥65 years.**

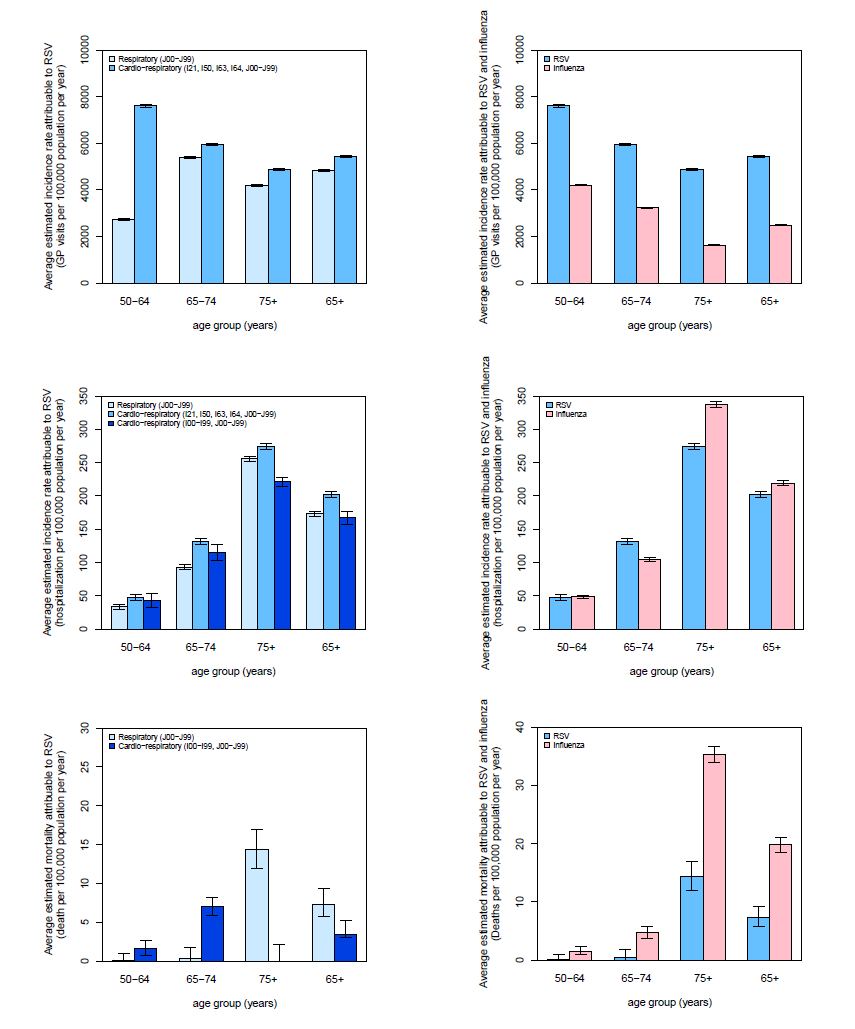

### **Supplementary Figure 3. Average estimated incidence rate of general practitioner visits, hospitalizations, and deaths attributable to respiratory syncytial virus or influenza infection. Average yearly incidence rates of general practitioner (GP) visits, hospitalizations, and deaths per 100,000 population attributable to respiratory syncytial virus (RSV) or influenza infection were estimated from cyclic Poisson regression models by age group from June 2013 to February 2020 (GP visits) or June 2010 to February 2020 (hospitalizations and deaths). The incidence rates for individuals aged ≥65 years were estimated independently, resulting in numbers of cases that differed from the sum of the groups aged 65–74 years and ≥75 years. Analysis were performed on respiratory causes (ICD-10 codes: J00−J99), cardiorespiratory causes (ICD-10 codes: I00−I99, J00−J99), or narrowly defined cardiorespiratory causes (ICD-10 codes: I21, I50, I63 [cerebral infarction]; ICD-10 codes: I64, J00−J99 [respiratory system]) as the primary diagnosis or primary and secondary diagnoses.**

### **Supplementary Table 1. List of ICD-10 codes for outcomes related to respiratory syncytial virus or influenza infection**

| **ICD-10 code** | **Label** |
| --- | --- |
| **Cardiorespiratory codes** | |
| I00–I99 | Circulatory system |
| J00–J99 | Respiratory system |
| **Narrowly defined cardiorespiratory codes** | |
| I21 | Acute myocardial infarction |
| I50 | Heart failure |
| I63 | Cerebral infarction |
| I64 | Stroke, not specified as hemorrhage or infarction |
| J00–J99 | Respiratory system |
| **Respiratory** | |
| J00–J99 | Respiratory system |
| **RSV codes** | |
| J12.1 | RSV pneumonia |
| J20.5 | Acute bronchitis due to RSV |
| J21.0 | Acute bronchiolitis due to RSV |
| J21.9a | Acute bronchiolitis, unspecified |
| B974b | RSV as the cause of diseases classified to other chapters |
| **Influenza codes** | |
| J09 | Influenza due to identified zoonotic or pandemic influenza virus |
| J10 | Influenza due to identified seasonal influenza virus |
| J11 | Influenza, virus not identified |

Abbreviations: ICD-10, International Classification of Diseases, 10^th^ Revision; RSV, respiratory syncytial virus

**^a^** Used only for children aged <2 years (primary objective – cyclic Poisson regression models).

^b^ Used only for adults (secondary objective – description of hospitalizations characteristics and associated costs)

### **Supplementary Table 2. Choice of time lags used in the model**

|  | **Lag for bronchiolitis (in weeks) – Lag for influenza (in weeks)** | | | |
| --- | --- | --- | --- | --- |
|  | 50–64 years | 65–74 years | ≥75 years | ≥65 years |
| **Outcomes** |  |  |  |  |
| GP visits | 1–0 | 1–0 | 1–0 | 1-0 |
| Hospitalizations | 1–0 | 1–0 | 1–0 | 1-0 |
| Deaths | 2–1 | 2–1 | 3–2 | 2-1 for 65-74 years  / 3-2 for ≥75 years |

### **Supplementary Table 3. Proportion of general practitioner visits, hospitalizations, and deaths attributable to respiratory syncytial virus and influenza among all the respiratory or cardiorespiratory cases, 2010–2020**

|  |  | **Proportion^a^ attributable to RSV (%)** | | **Proportion^a^ attributable to influenza (%)** | | |
| --- | --- | --- | --- | --- | --- | --- |
|  | **Age group**  **(years)** | **Overall study period** | **Over winter seasons only [November–March]** | | **Overall study**  **period** | **Over winter seasons only**  **[November–March]** |
| **GP VISITS (2013–2020)** | |  |  |  | |  |
| **Respiratory disease**  **(ICD-10 codes: J00−J99)** | **50–64** | 3.62 | 6.49 | 5.10 | | 9.16 |
|  | **65–74** | 6.29 | 11.50 | 3.42 | | 6.25 |
|  | **≥75** | 5.27 | 9.91 | 2.40 | | 4.52 |
|  | **≥65** | 5.84 | 10.81 | 2.97 | | 5.50 |
| **Cardiorespiratory disease**  **(ICD-10 codes: I21, I50, I63, I64, J00−J99)** | **50–64** | 9.40 | 17.14 | 5.20 | | 9.47 |
|  | **65–74** | 6.15 | 11.45 | 3.30 | | 6.15 |
|  | **≥75** | 4.79 | 9.29 | 1.61 | | 3.12 |
|  | **≥65** | 5.50 | 10.44 | 2.50 | | 4.74 |
| **HOSPITALIZATIONS (2010–2020)** | | |  |  | |  |
| **Respiratory disease**  **(ICD-10 codes: J00−J99)** | **50–64** | 3.77 | 7.61 | 4.93 | | 9.93 |
|  | **65–74** | 5.84 | 11.44 | 5.89 | | 11.55 |
|  | **≥75** | 6.06 | 11.34 | 6.37 | | 11.92 |
|  | **≥65** | 6.00 | 11.37 | 6.24 | | 11.82 |
| **Narrowly defined cardiorespiratory disease**  **(ICD-10 codes: I21, I50, I63, I64, J00−J99)** | **50–64** | 3.13 | 6.54 | 3.15 | | 6.59 |
|  | **65–74** | 4.34 | 8.92 | 3.47 | | 7.14 |
|  | **≥75** | 2.91 | 5.82 | 3.57 | | 7.16 |
|  | **≥65** | 3.26 | 6.57 | 3.55 | | 7.15 |
| **Cardiorespiratory disease**  **(ICD-10 codes: I00-I99, J00−J99)** | **50–64** | 1.20 | 2.50 | 1.01 | | 2.11 |
|  | **65–74** | 1.66 | 3.47 | 1.17 | | 2.44 |
|  | **≥75** | 1.51 | 3.12 | 2.44 | | 5.05 |
|  | **≥65** | 1.56 | 3.23 | 2.02 | | 4.20 |
| **DEATHS (2010–2020)** | | |  |  | |  |
| **Respiratory disease**  **(ICD-10 codes: J00−J99)** | **50–64** | 0.47 | 0.90 | 8.20 | | 15.59 |
|  | **65–74** | 0.55 | 1.05 | 7.40 | | 14.14 |
|  | **≥75** | 2.71 | 4.94 | 6.67 | | 12.14 |
|  | **≥65** | 2.47 | 4.52 | 6.75 | | 12.36 |
| **Cardiorespiratory disease**  **(ICD-10 codes: I00−I99, J00−J99)** | **50–64** | 1.83 | 3.81 | 2.94 | | 6.11 |
|  | **65–74** | 2.45 | 5.06 | 2.86 | | 5.92 |
|  | **≥75** | 0.00 | 0.00 | 2.87 | | 5.76 |
|  | **≥65** | 0.27 | 0.54 | 2.87 | | 5.78 |

Abbreviations: ICD-10, International Classification of Diseases, 10^th^ Revision; GP, general practitioner; RSV, respiratory syncytial virus

**^a^** Proportion of the estimated number of GP visits, hospitalizations, and deaths attributable to RSV and influenza infection among the total number of these events coded with respiratory (ICD-10 codes: J00–J99) and cardiorespiratory (circulatory and respiratory system ICD-10 codes: I00–I99, J00–J99; respiratory system ICD-10 codes: I21, I50, I63, I64, J00-J99) primary diagnosis codes. This proportion is calculated for the total period (i.e., 2013–2020 or 2010–2020) and restricted to the cold seasons (i.e., November–March) over the total period.

Supplementary Table 4. Variation in the primary diagnosis ICD-10 codes necessitating hospitalization which have a respiratory syncytial virus ICD-10 code (J121, J205, J210, B974) as the secondary diagnosis code, 2015–2019

| **ICD-10 code: definition** | **RSV hospitalizations** **N (%)** |
| --- | --- |
| **Top 10 primary respiratory and cardiac diagnosis codes for patients hospitalized with a secondary diagnosis of RSV** | 6,480 (100%) |
| J440: Chronic obstructive pulmonary disease with acute lower respiratory tract infection | 814 (12.6) |
| I501: Left ventricular failure | 360 (5.6) |
| I500: Congestive heart failure | 343 (5.3) |
| J9600: Acute respiratory failure type I (hypoxic) | 243 (3.8) |
| J9601: Acute respiratory failure type II (hypercapnic) | 195 (3.0) |
| J441: Chronic obstructive pulmonary disease with acute episodes, unspecified | 166 (2.6) |
| J960: Acute respiratory failure | 161 (2.5) |
| J13: Pneumonia caused by *Stretococcus pneumoniae* | 129 (2.0) |
| J46: Status asthmaticus | 92 (1.4) |
| J159: Bacterial pneumonia, unspecified | 82 (1.3) |
| **Type of primary diagnosis** | |
| Respiratory | 7,838 (71.2) |
| Cardiac | 1,219 (11.1) |
| Other | 1,948 (17.7) |
| **Top 10 non-respiratory and non-cardiac primary diagnosis codes for patients hospitalized with a secondary diagnosis of RSV** | 1,948 (100%) |
| N10: Acute tubulointerstitial nephritis | 55 (2.8) |
| Z511: Chemotherapy session for tumor eradication | 50 (2.6) |
| C900: Multiple myeloma | 49 (2.5) |
| T796: Traumatic muscle ischemia | 49 (2.5) |
| D611: Drug-induced bone marrow aplasia | 48 (2.5) |
| Z515: Palliative care | 45 (2.3) |
| C920: Acute myeloblastic leukemia | 33 (1.7) |
| N410: Acute prostatitis | 28 (1.4) |
| N179: Acute renal failure, unspecified | 24 (1.2) |
| C833: Diffuse large B-cell lymphoma | 23 (1.2) |

Abbreviations: ICD-10, International Classification of Diseases, 10^th^ Revision; RSV, respiratory syncytial virus
